# Association between endotypes of prematurity and pharmacological closure of patent ductus arteriosus: A systematic review and meta-analysis

**DOI:** 10.1101/2022.10.24.22281438

**Authors:** Gema E Gonzalez-Luis, Moreyba Borges-Lujan, Eduardo Villamor

## Abstract

Endotypes leading to very and extremely preterm birth are clustered into two groups: infection/inflammation and dysfunctional placentation. We conducted a systematic review of observational studies exploring the association between these two endotypes and the pharmacological closure of patent ductus arteriosus (PDA) induced by cyclooxygenase (COX) inhibitors. Chorioamnionitis represented the infectious-inflammatory endotype, while dysfunctional placentation proxies were hypertensive disorders of pregnancy (HDP) and small for gestational age (SGA) or intrauterine growth restriction. PubMed/Medline and Embase databases were searched. The random-effects odds ratio (OR) and 95% confidence interval (CI) were calculated for each association. We included 30 studies (12639 infants). Meta-analysis showed a significant association between exposure to HDP and increased rate of pharmacological closure of PDA (17 studies, OR 1.41, 95% CI 1.10-1.81, p=0.006). In contrast, neither chorioamnionitis (13 studies, OR 0.75, 95% CI 0.47-1.18, p=0.211) nor SGA (17 studies, OR 1.20, 95% CI 0.96-1.50, p=0.115) were significantly associated with the response to therapy. Subgroup analyses showed that the higher response to COX inhibitors in the HDP group was significant for indomethacin (OR 1.568, 95% CI 1.147-2.141, p=0.005) but not for ibuprofen (OR 1.107, 95% CI 0.248-4.392, p=0.894) or for the studies using both drugs (OR 1.280, 95% CI 0.935-1.751, p=0.124). However, meta-regression showed that this difference between the drugs was not statistically significant (p=404). In conclusion, our data suggest that the pathologic condition that triggers prematurity may alter the response to pharmacological treatment of PDA. The DA of infants exposed to HDP appears to be more responsive to COX inhibitors.

## 1. Introduction

The ductus arteriosus (DA) is a fetal vessel connecting the pulmonary artery to the descending aorta, which is of major functional importance for the integrity of the fetal circulation (1-6). Morphological and functional maturation of the DA during fetal life prepares the vessel for functional and then anatomical postnatal closure. This closure is a key event in the transition to extrauterine life. However, when preterm birth interrupts physiological maturation, infants are exposed to patent ductus DA (PDA) due to underdeveloped ductal closure mechanisms (1-6).

PDA in very and extremely preterm infants (i.e. with gestational age less than 32 weeks) is a persistent dilemma for neonatal medicine, as well as a potential morbidity and mortality contributor when hemodynamically significant (1, 7, 8). Although the therapeutic approach has changed and continues to change over the years, the classical pharmacological treatment of PDA is based on cyclooxygenase (COX) inhibitors, such as indomethacin and ibuprofen (9-12). Paracetamol has been added to these drugs in recent years (9).

Low gestational age (GA) is the main risk factor for both failure of spontaneous and pharmacologic closure of the DA (2, 13-15) but there is a growing recognition that the pathologic conditions that trigger preterm birth may play a relevant role in in the incidence of PDA, as well as in the response to COX inhibitors (16-20). The two main pathophysiologic pathways, or endotypes, leading to very/extremely preterm birth are 1) infection/inflammation and 2) dysfunctional placentation (21-24). Previous studies by our group and other investigators have systematically reviewed the association between these two endotypes and a number of complications of prematurity including PDA (15, 22, 25-31). In these meta-analyses, the infectious-inflammatory endotype was represented by chorioamnionitis, while the placental dysfunction endotype was represented by hypertensive disorders of pregnancy (HDP) and fetal growth restriction (15, 22, 25-31).

Individual studies suggest that prenatal conditions such as chorioamnionitis (16, 17) or HDP (18-20) may affect the therapeutic response to COX inhibitors. However, these findings have not been systematically reviewed. Our aim in the present study is to fill this gap in the literature by conducting a systematic review and meta-analysis on the association between endotype of prematurity and pharmacological closure of PDA in very and extremely preterm infants.

## 2. Methods

The methodology of the present study is based on that used in our previous meta-analyses on the association of several risk factors and the incidence of PDA and/or the response to pharmacological treatment of PDA (26, 27, 32-34). The study was performed and reported according to the preferred reporting items for systematic reviews and meta-analyses (PRISMA) and meta-analysis of observational studies in epidemiology (MOOSE) guidelines [15]. Review protocol was registered in the PROSPERO international register of systematic reviews (ID= CRD42018095509). The Population, Exposure, Comparison and Outcome (PECO) question was: Do very/extremely preterm infants (P) exposed to chorioamnionitis, HDP, or growth restriction during pregnancy (E) have a different rate of PDA closure in response to treatment with COX inhibitors (O) than infants with no history of exposure (C)?

### Sources and search strategy

A comprehensive literature search was undertaken using the PubMed and EMBASE databases. The search terms involved various combinations of the following key words: *ductus arteriosus, patent ductus arteriosus, PDA, Patency of the Ductus Arteriosus, Ductus Botalli, treatment, pharmacologic(al) closure, indomethacin, ibuprofen, paracetamol, acetaminophen, cyclooxygenase, COX, chorioamnionitis, intrauterine infection, intrauterine inflammation, antenatal infection, antenatal inflammation, preeclampsia, IUGR, growth restriction, growth retardation, restricted growth, fetal growth, fetus growth, placental dysfunction, placental insufficiency, chronic hypoxia, chronic hypoxemia, small for gestational age, small for date, SGA, gestational hypertension, maternal hypertension, hellp syndrome, hypertensive disorders, toxemia, hypertensive disorders of pregnancy*. No language limit was applied. The literature search was updated up to March 2022. Narrative reviews, systematic reviews, case reports, letters, editorials, and commentaries were excluded, but read to identify potential additional studies. Additional strategies to identify studies included manual review of reference lists from key articles that fulfilled our eligibility criteria, use of “related articles” feature in PubMed, and use of the “cited by” tool in Web of Science and Google scholar.

### Study selection and definitions

Studies were included if they had a prospective or retrospective design, examined infants with GA below 32 weeks and reported primary data that could be used to measure the association between pharmacological closure of PDA and exposure to chorioamnionitis (clinical or histological), HDP (including pregnancy-induced hypertension, preeclampsia and eclampsia) or fetal growth restriction. As we did in our previous meta-analyses, we accepted small for gestational age (SGA) as a proxy for fetal growth restriction (22, 27). Regarding response to drug treatment, when a study reported on several treatment courses, only the final response was taken into account. To identify relevant studies, two reviewers (GG-L and MB-L) independently screened the results of the searches and applied inclusion criteria using a structured form. Discrepancies were resolved by the third reviewer (EV).

### Data Extraction and Assessment of Study Quality

Two investigators (GG-L and MB-L) extracted data on study design, demographics, and response to treatment. Another investigator (EV) checked the data extraction for completeness and accuracy. Methodological quality was assessed using the Newcastle-Ottawa Scale (NOS) for cohort studies [15]. This scale assigns a maximum of 9 points (4 for selection, 2 for comparability, and 3 for outcome). NOS scores≥ 7 were considered high-quality studies (low risk of bias), and scores of 5 to 6 denoted moderate quality (moderate risk of bias) [15].

### Statistical Analysis

Studies were combined and analyzed using comprehensive meta-analysis V3.0 software (Biostat Inc., Englewood, NJ, USA). The odds ratio (OR) with 95% confidence interval (CI) was calculated with a random-effects model and subgroups were combined with a mixed-effects model (35). Statistical heterogeneity was assessed by Cochran’s *Q* statistic and by the *I*^*2*^ statistic. I^2^ was interpreted on the basis of Higgins and Thompson criteria, where 25%, 50%, and 75% correspond to low, moderate, and high heterogeneity, respectively (36). Potential sources of heterogeneity were assessed through subgroup analysis and/or random effects (method of moments) univariate meta-regression analysis as previously described [16, 17]. For both categorical and continuous covariates, the R^2^ analog, defined as the total between-study variance explained by the moderator, was calculated based on the meta-regression matrix. Predefined sources of heterogeneity included the following characteristics of cohorts: definition of exposure (clinical or histological chorioamnionitis, HDP definition, and growth restriction or SGA definition), mean or median GA, median year of birth, and drug used for PDA treatment. We used the Egger’s regression test and funnel plots to assess publication bias. Subgroup analyses, meta-regression, and publication bias assessment were performed only when there were at least ten studies in the meta-analysis. A probability value of less than 0.05 (0.10 for heterogeneity) was considered statistically significant.

## 3. Results

### Description of studies and quality assessment

The flow diagram of the search process is shown in Figure S1. Of 964 potentially relevant studies, 30 (including 12639 infants) were included (13, 16-20, 37-60). Their characteristics are summarized in Table S1. Focusing on exposure, 13 studies provided data on chorioamnionitis, 17 on HDP, and 17 on SGA. We found no studies that evaluated IUGR (i.e., assessment of growth during fetal period). Nineteen studies reported on indomethacin (16-20, 37-50), five on ibuprofen (51-55) and in six studies both drugs were used (13, 56-60). The quality score of each study according to the Newcastle-Ottawa Scale is depicted in Table S1. All studies received at least 7 points indicating a low risk of bias.

### Meta-analysis

Meta-analysis could not demonstrate a significant association between chorioamnionitis and response to pharmacological treatment of PDA (OR 0.746, 95% CI 0.472-1.181, p=0.211) (Figure 1). This lack of significant effect of chorioamnionitis was observed for both clinical (OR 0.838, 95% CI 0.492-1.428, p=0.516) and histological chorioamnionitis (OR 0.536, 95% CI 0.217-1.319, p=0.175). In contrast, meta-analysis showed a significant association between exposure to HDP and pharmacological closure of PDA (OR 1.413, 95% CI 1.102-1.811, p=0.006) (Figure 2). Subgroup analysis showed that this significant association was maintained in the any HDP group (OR 1.392, 95% CI 1.004-1.931, p=0.006) but not in the preeclampsia group (OR 1.441, 95% CI 0.984-2.109, p=0.061). Finally, the meta-analysis could not demonstrate a significant association between being SGA and response to pharmacological treatment of PDA (OR 1.197, 95% CI 0.957-1.497, p=0.115) (Figure 3). This lack of significant association was consistent for all SGA definitions (Figure 3). Neither visual inspection nor Egger’s test suggested the presence of publication or selection bias for none of the three meta-analyses (Figure S2).

**Figure 1.**
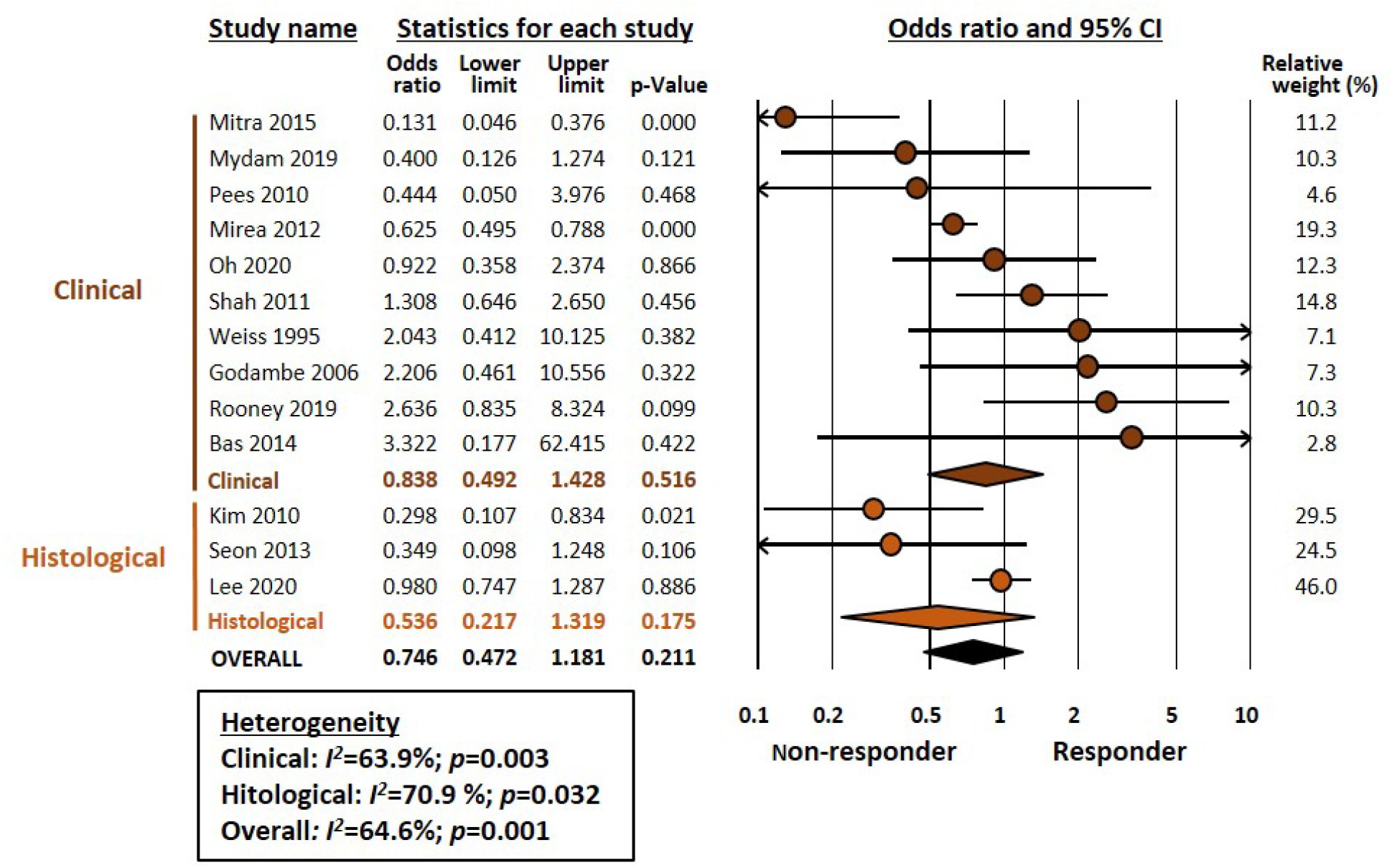
Meta-analysis on the association between chorioamnionitis and pharmacological closure of patent ductus arteriosus.

**Figure 2.**
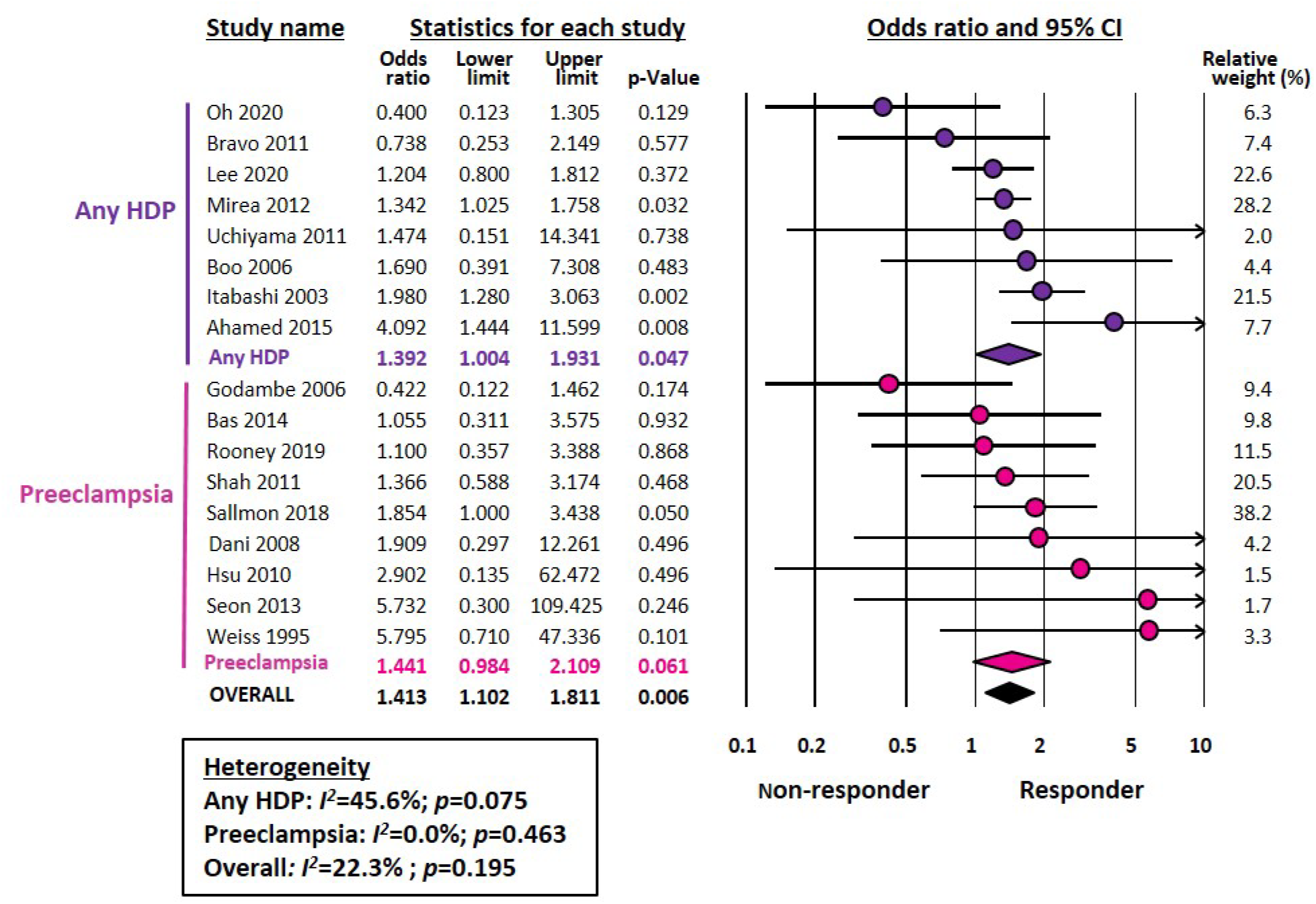
Meta-analysis on the association between hypertensive disorders of pregnancy (HDP) and pharmacological closure of patent ductus arteriosus.

**Figure 3.**
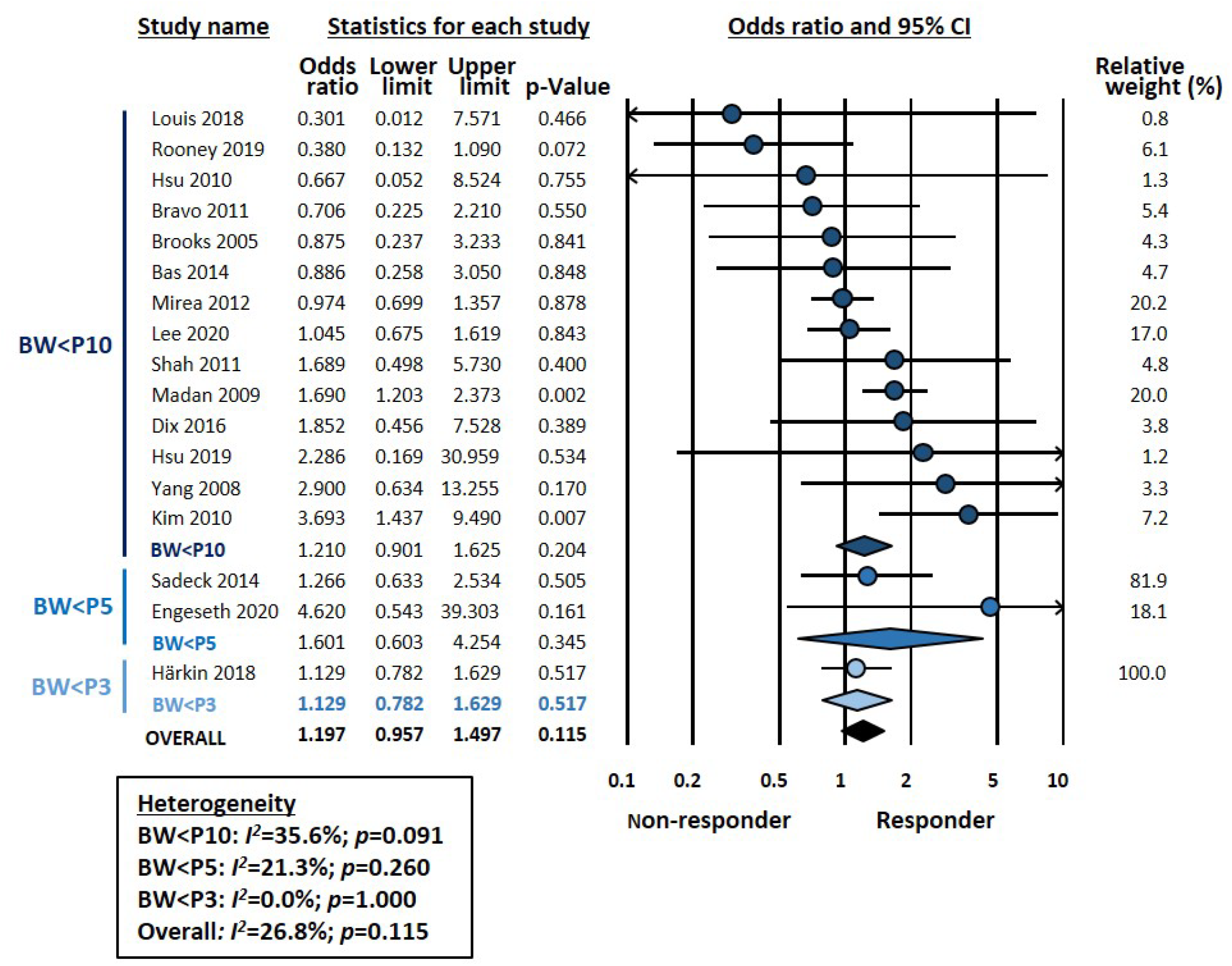
Meta-analysis on the association between being small for gestational age (SGA) and pharmacological closure of patent ductus arteriosus.

### Subgroup analysis and meta-regression

In addition to the subgroup analysis based on the definition of the different exposures (Figures 1-3), we conducted a second analysis based on the drug used to treat PDA. As shown in Table 1and Figure S3, the association between HDP and pharmacologic closure of PDA was significant only for indomethacin but not for ibuprofen or for studies using either indomethacin or ibuprofen. However, meta-regression showed that this difference between the drugs was not statistically significant (P=404, Figure S3). In the chorioamnionitis and SGA meta-analyses, no significant association with any of the drugs was detected (Table 1, Figure S3).

**Table 1.** Subgroup analysis based on drug used for the treatment of patent ductus arteriosus

| Meta-analysis | Drug | K | OR | 95% CI |  | P | Heterogeneity |  | Meta-regression P-value |
| --- | --- | --- | --- | --- | --- | --- | --- | --- | --- |
|  |  |  |  | Lower limit | Upper limit |  | I<sup>2</sup> | P |  |
| Hypertensive disorders of pregnancy | Any | 4 | 1.280 | 0.935 | 1.751 | 0.124 | 0.0 | 0.460 | 0.404 |
|  | Ibuprofen | 3 | 1.107 | 0.248 | 4.932 | 0.894 | 48.4 | 0.144 |  |
|  | Indomethacin | 10 | 1.568 | 1.147 | 2.141 | 0.005 | 24.4 | 0.219 |  |
| Chorioamnionitis | Any | 2 | 0.991 | 0.755 | 1.299 | 0.945 | 0.0 | 0.417 | 0.673 |
|  | Ibuprofen | 3 | 0.627 | 0.306 | 1.284 | 0.202 | 0.0 | 0.462 |  |
|  | Indomethacin | 8 | 0.725 | 0.399 | 1.319 | 0.292 | 72.7 | 0.001 |  |
| Small for gestational age | Any | 5 | 1.082 | 0.844 | 1.388 | 0.533 | 0.0 | 0.923 | 0.148 |
|  | Ibuprofen | 1 | 2.286 | 0.169 | 30.959 | 0.534 | 0.0 | 1.000 |  |
|  | Indomethacin | 11 | 1.372 | 0.914 | 2.059 | 0.127 | 48.4 | 0.036 |  |

CI: confidence interval, K: number of studies, OR: odds ratio.

We also analyzed by meta-regression how the median year of the cohort (Table 2, Figure S4) or the gestational age of the included infants (Table 2, Figure S5) could influence the effect size of the different meta-analyses. The only significant finding from these meta-regressions (Table 2, Figure S4) was the correlation between the median year of the cohort and the effect size of the association between SGA and drug response (p=0.008). As the cohort became more modern, the effect size of the association significantly decreased (Table 2, Figure S4).

**Table 2.** Meta-regression analysis of continuous covariates

| Meta-analysis | Covariate | K | Coefficient | 95% CI |  | P-value | R<sup>2</sup> analog |
| --- | --- | --- | --- | --- | --- | --- | --- |
|  |  |  |  | Lower limit | Upper limit |  |  |
| Chorioamnionitis | GA cohort (weeks) | 13 | 0.478 | -0.125 | 1.081 | 0.120 | 0.00 |
|  | Median year of cohort | 13 | -0.035 | -0.130 | 0.060 | 0.469 | 0.00 |
| Hypertensive disorders of pregnancy | GA cohort (weeks) | 17 | 0.027 | -0.270 | 0.324 | 0.859 | 0.00 |
|  | Median year of cohort | 17 | -0.037 | -0.076 | 0.002 | 0.063 | 0.61 |
| Small for gestational age | GA cohort (weeks) | 17 | -0.095 | -0.264 | 0.074 | 0.269 | 0.00 |
|  | Median year of cohort | 17 | -0.049 | -0.093 | -0.004 | 0.032 | 0.71 |

CI: confidence interval, GA: gestational age, K: number of studies, OR: odds ratio.

## 4. Discussion

To the best of our knowledge, the present study is the first systematic review and meta-analysis examining the effect of endotype of prematurity on the response to pharmacological treatment of PDA in very or extremely preterm infants. We analyzed three pathological conditions (chorioamnionitis, HDP, and SGA) and observed that HDP was associated with a higher rate of pharmacologic closure of PDA. Moreover, subgroup analysis showed that the association between HDP and response to treatment was particularly significant for indomethacin. However, the relatively small number of studies and the moderate degree of heterogeneity that was present in some of the meta-analyses limit our results.

As mentioned in the introduction, the different endotypes of prematurity are not only the trigger for preterm birth but also induce a different pathophysiological environment for the development of fetal organs and systems. Thus, the infectious-inflammatory endotype is characterized by the development of a systemic inflammatory response with elevated cytokine levels (21, 23, 24). In turn, the placental dysfunction endotype is characterized by chronic hypoxia and imbalance of pro- and anti-angiogenic factors (21-24). Both pathophysiological pathways could plausibly affect the normal development of the DA resulting in delay or failure of its spontaneous or pharmacological closure.

Peri- and postnatal infection is classically considered a key risk factor for PDA (16, 17, 61-64). Two previous meta-analyses showed a significant association between chorioamnionitis and risk of developing PDA in very preterm infants (26, 31). However, a significant proportion of the risk appears to be associated with the lower GA of infants with chorioamnionitis compared to those not exposed to the insult (26). Interestingly, the presence of fetal inflammatory response (i.e. funisitis) did not increase the risk of PDA when compared to chorioamnionitis in the absence of funisitis (26). Although intrauterine infection is associated with increased COX expression and prostaglandin production (17, 61, 63, 65), the present meta-analysis could not demonstrate an association between chorioamnionitis and response to COX inhibitors.

The only antenatal pathology for which we have found an association with pharmacological closure of the DA was HDP. That HDP may affect the incidence of PDA has been demonstrated in several cohort studies (66-68). However, a recent meta-analysis, including eight studies, found no significant association between HDP and the risk of developing PDA (15). There are several potential explanations for the higher rate of spontaneous and pharmacologic ductal closure in preterm infants with intrauterine exposure to HDP. Women with HDP are likely to deliver before the onset of natural labor and therefore the production of prostaglandins is lower than that of pregnant women in labor (69). In addition, preterm infants born due to HDP tend to have higher gestational ages than those born due to chorioamnionitis (22). This leads to higher clinical stability and lower incidence of respiratory complications (22, 23). Finally, exposure to HDP does not induce the inflammatory response that is induced by chorioamnionitis. It should be noted that extremely preterm birth is by definition a pathological condition. Therefore, there is no “healthy control group” to compare with. The positive effects of HDP on pharmacological closure of the DA may reflect not the direct action of HDP but the absence, or at least attenuated presence, of an infectious process.

Subgroup analysis showed that the association between HDP and higher rate of pharmacologic ductal closure was particularly significant for indomethacin. However, meta-regression showed that the difference between the drugs was not statistically significant. Interestingly, Louis et al. reported that the rate of ductal closure induced by prophylactic indomethacin was significantly higher in the offspring of HDP mothers (70). With our present results, we can only speculate on the potential mechanisms responsible for this difference between indomethacin and ibuprofen. A Cochrane meta-analysis concluded that ibuprofen was as effective as indomethacin for PDA closure, whereas the former reduced the risk of necrotizing enterocolitis and transient renal insufficiency (71). The choice of indomethacin or ibuprofen to treat PDA is highly variable among neonatologists and is often influenced by non-clinical factors such as difficulties with drug supply (10-12). Nevertheless, there are pharmacokinetic, pharmacodynamic and pharmacogenetic differences between ibuprofen and indomethacin that may account for a different response in particular subgroups of preterm infants (72-74).

The third condition in which we have analyzed pharmacological ductal closure is fetal growth restriction. It should be noted that these results were limited because all included studies reported data on SGA and not IUGR. Although the terms SGA and IUGR are often used synonymously, SGA is a statistical definition based on BW, with the 10th percentile as the most frequently used cut-off (75-77). Therefore, the term SGA also encompasses constitutionally small infants without growth restriction (75-77). On the other hand, infants with pathologic growth restriction may have a BW above the 10th percentile (75-77). In a recent meta-analysis we investigated the association between SGA/IUGR and incidence of PDA. Although we observed a negative association, i.e. the rate of any PDA was lower in the growth-restricted group, this association only involved the subgroup in which SGA was defined using the 10th percentile threshold (27). Moreover, when examining the subgroup of studies that used a definition for growth restriction that went beyond BW for GA (i.e. assessment of fetal growth or presence of abnormal Doppler), meta-analysis could not find a significant association with PDA (27). In addition, we could not find a significant association between SGA/IUGR and the development of hemodynamically significant PDA (27). In the present meta-analysis, we could only find studies focusing on SGA infants, mostly defined by the 10th percentile cutoff. Although very limited by this fact and the low number of studies, our data do not suggest that being SGA affects the rate of pharmacological closure of PDA.

Neonatology is a medical specialty in constant evolution and this is also reflected in the diagnostic and therapeutic approach to PDA. Neonatologists have been debating for decades what is a hemodynamically significant PDA, what are the health consequences of the presence of a ductal shunt for the preterm newborn and when and how PDA should be treated (1, 7, 8). As a result of this debate, there has been a transition from advocating that all PDA should be treated, with an intermediate step in which only targeted group were treated, to a current trend of therapeutic nihilism. That is, the consideration of PDA in very preterm infants as a “physiological” condition whose treatment produces more harm than benefit (1, 7, 8). This may influence our results as the number of infants treated has probably decreased with the passing of time. We have analyzed by meta-regression whether the association between endotype of prematurity and pharmacological closure of the DA has changed over the years. We have found that as cohorts become more contemporary, the response to COX inhibitors in SGA infants tends to decrease. The course of time does not seem to have affected pharmacological ductal closure in infants exposed to chorioamnionitis or HDP. However, it should be noted that our meta-regression analysis is based on few studies. The minimum number of trials per covariate in meta-regression analyses required to minimize the risk of overfitting is unknown but it has been suggested a minimum of 10 studies per examined covariate (78, 79). The meta-regressions presented here include 15-16 studies and are therefore just above the minimum recommended threshold. Therefore, they have mainly an exploratory and hypothesis-generating value.

In conclusion, our data suggest that the pathological condition that triggers prematurity may alter not only the incidence of PDA but also the response to pharmacological treatment. Our meta-analysis further supports the growing evidence that not all preterm infants are identical even if they are of the same gestational age. Therefore, *one size fits all* is no longer an appropriate approach to perinatal medicine (23, 80-82). Personalized medicine requires an adequate characterization of endotypes and clinical phenotypes so that each infant can receive the therapeutic approach best suited to his or her individual characteristics.

## Data Availability

All data produced in the present study are available upon reasonable request to the authors

## Author Contributions

G G-L and M B-L selected studies for inclusion, collected data, contributed to the statistical analysis and interpretation of the results, collaborated in the preparation of the graphs and tables, and reviewed and revised the manuscript. EV conceptualized and designed the study, performed the search, supervised data collection, planned and performed the statistical analysis, and wrote the drafts of the manuscript. All authors approved the final manuscript as submitted.

## Acknowledgments

We thank Ana Martinez-Olaizola, MSc for her technical and administrative support.

## Data Availability Statement

All data relevant to the study are included in the article or uploaded as supplementary information. Additional data are available upon reasonable request.

