## Supplementary Figures and Tables for "Association between endotypes of prematurity and pharmacological closure of patent ductus arteriosus: A systematic review and meta-analysis"

### *Supplementary Material*

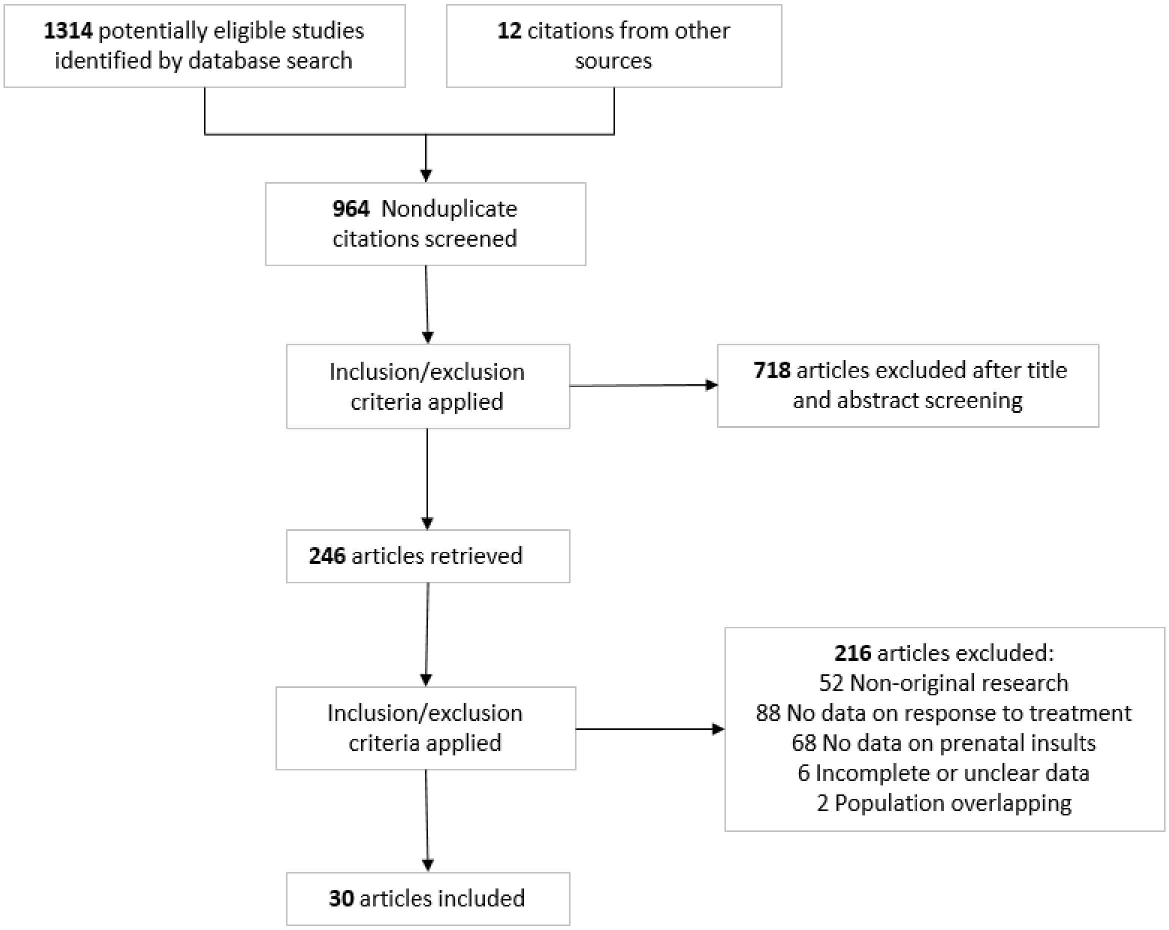


**Supplementary Figure 1.** Flow diagram of the systematic search

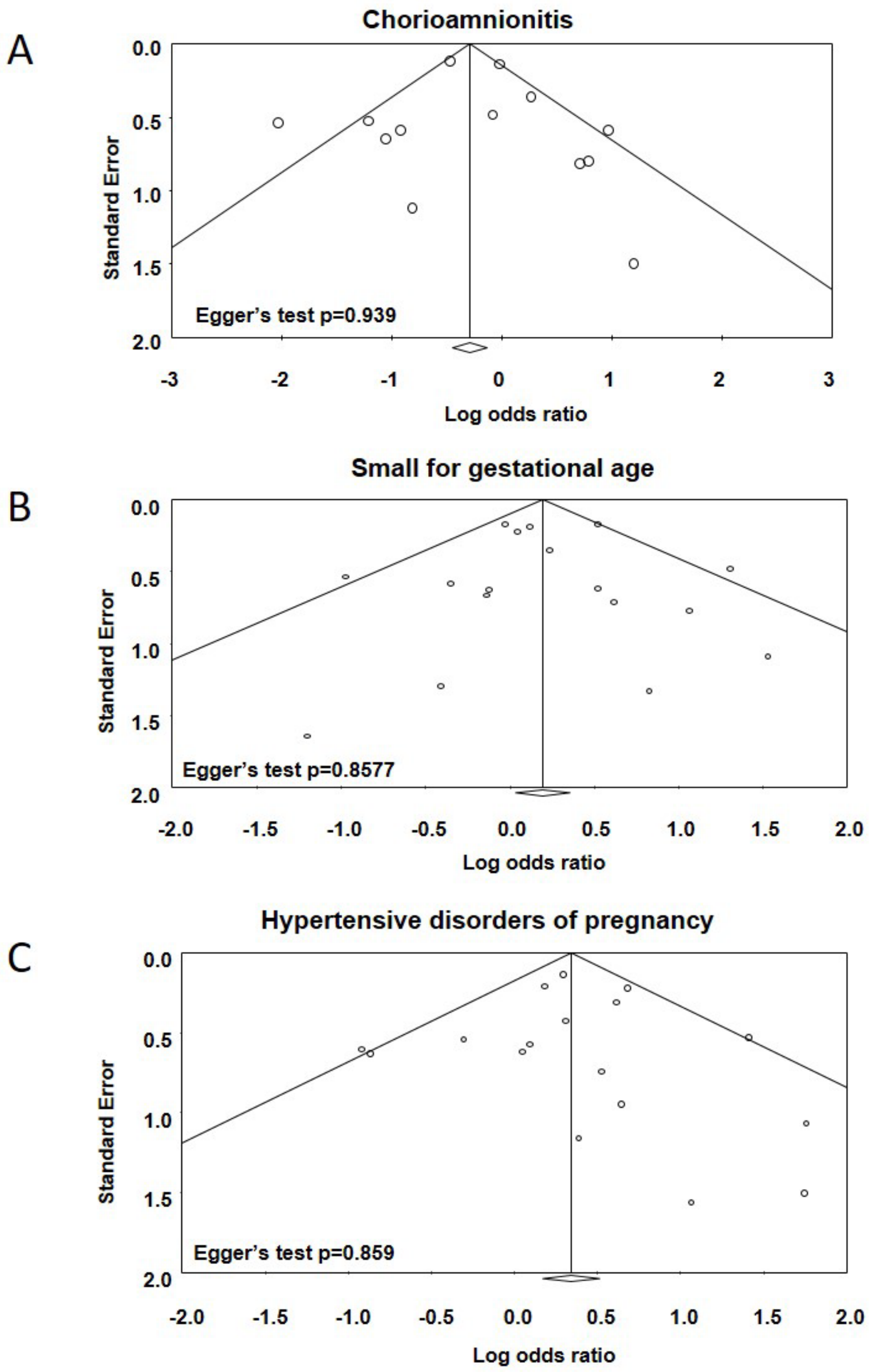


**Supplementary Figure 2.** Funnel plot for publication bias analysis for the studies included in the different meta-analyses.

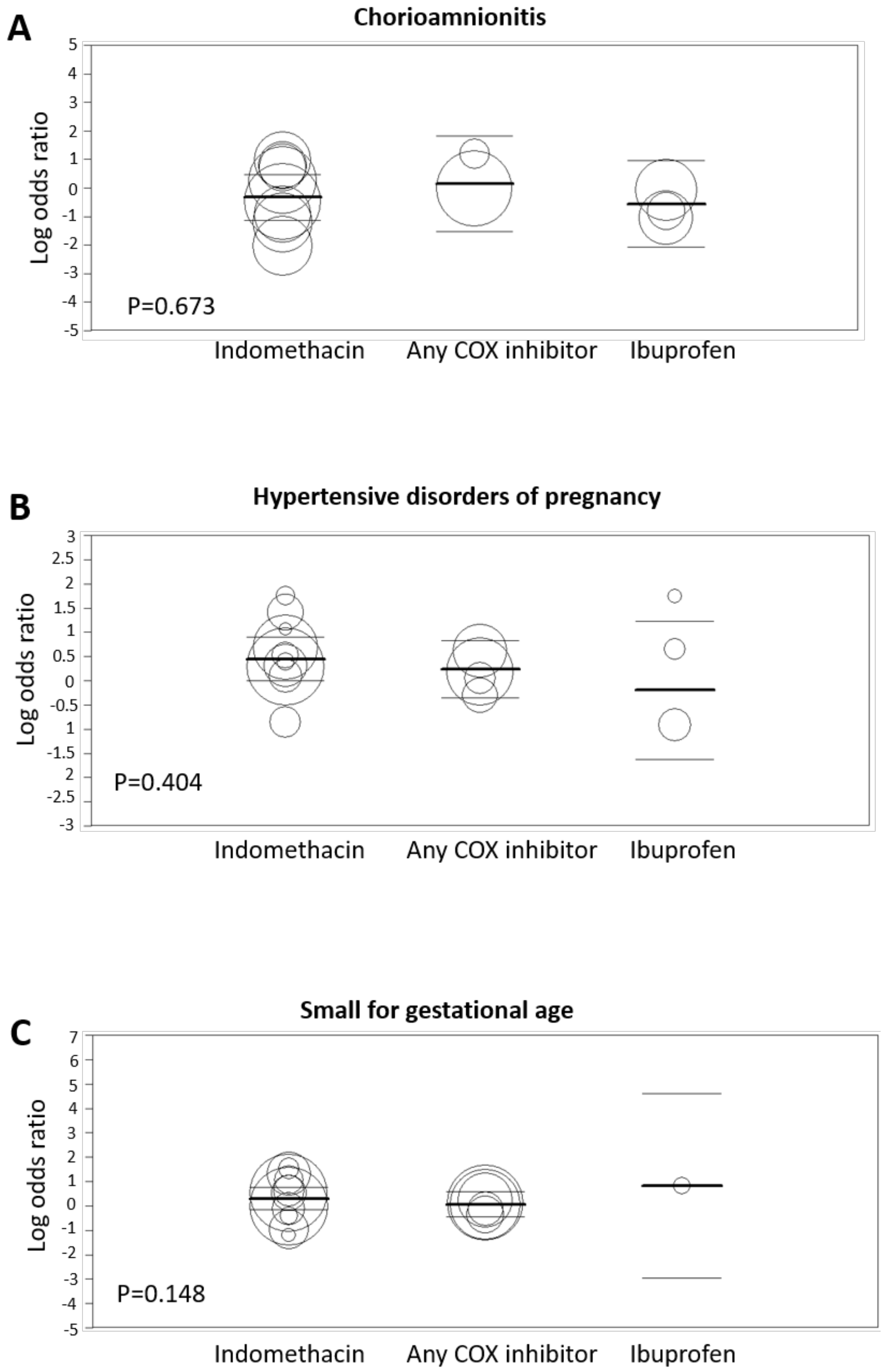


**Supplementary Figure 3.** Metaregression on the effect of the different cyclooxygenase (COX) inhibitors used in the studies.

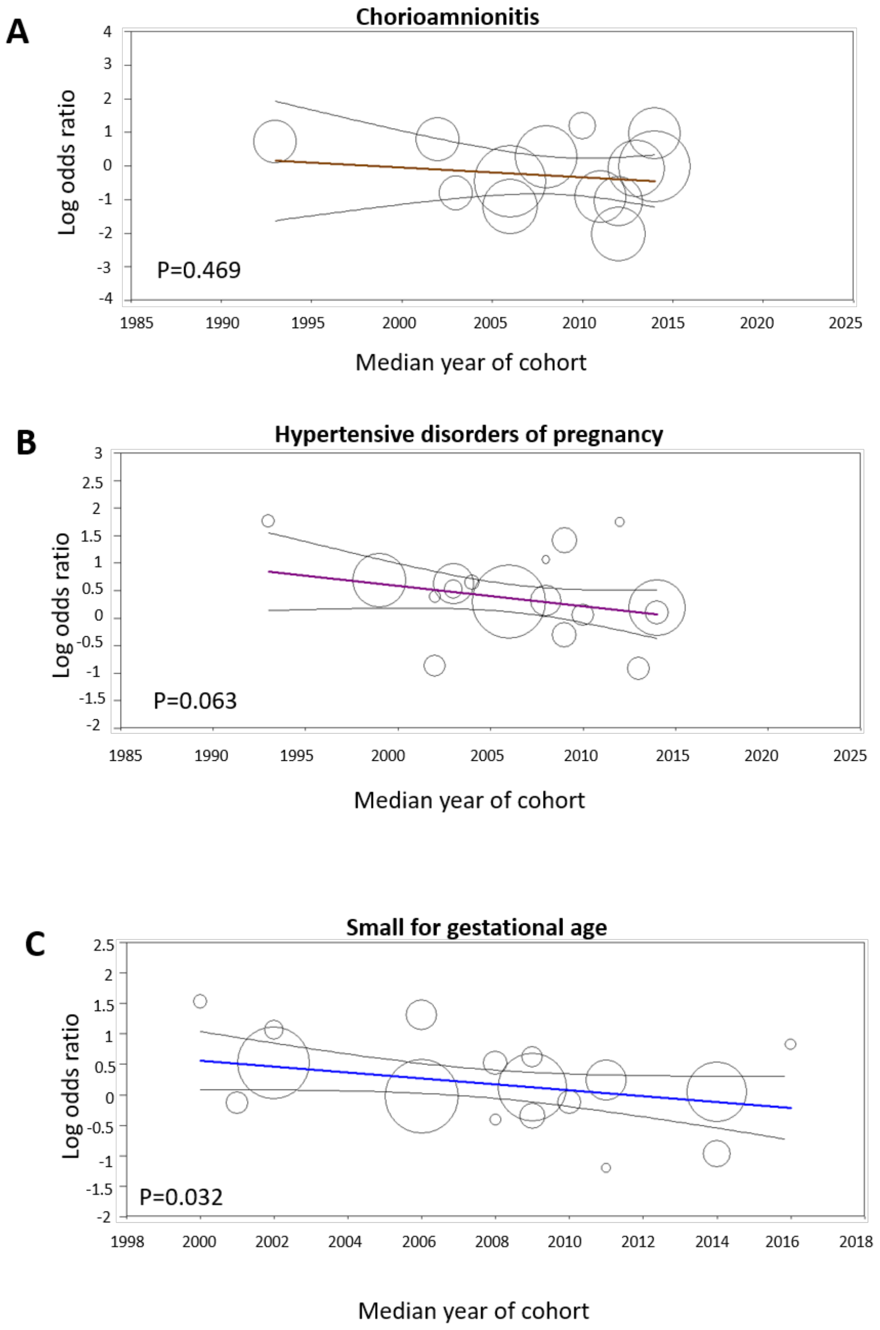


**Supplementary Figure 4.** Metaregression on the effect of the median year of the cohort in the different meta-analyses.

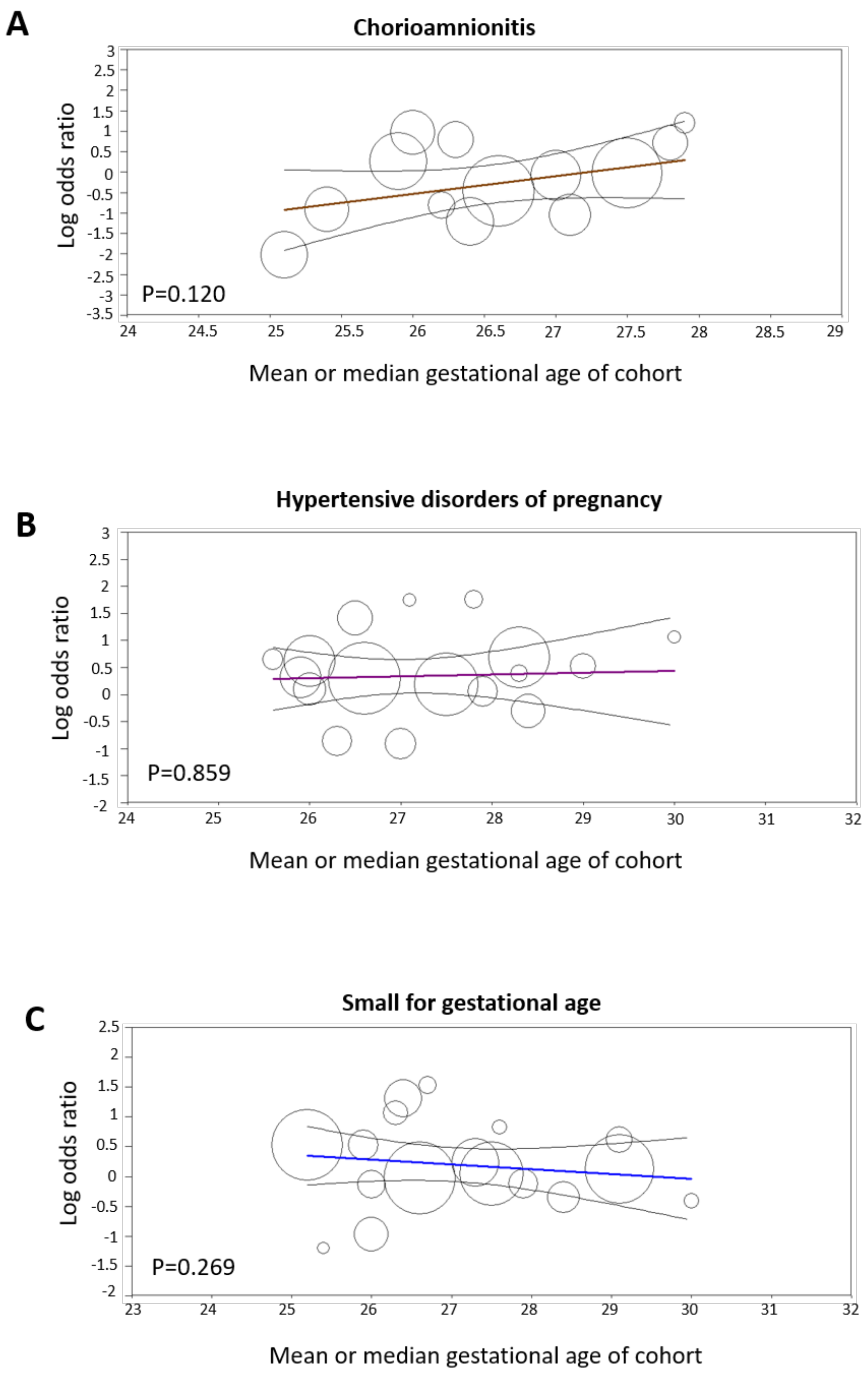


**Supplementary Figure 5.** Metaregression on the effect of gestational age of the cohort in the different meta-analyses.

**Supplementary Table 1.** Characteristics of the studies included in the meta-analyses.

| First author. year | Country | Design | Prospective? | Total infants | Centers | GA (mean or median) | BW mean/median | Exposure | Treatment | Selection | Comparability | Outcome/ Exposure | Total NOS |
|---|---|---|---|---|---|---|---|---|---|---|---|---|---|
| Ahamed 2015 (1) | USA | Cohort | No | 119 | 1 | 26.5 | 840.6 | HDP | INDO | 3 | 1 | 3 | 7 |
| Bas 2014 (2) | Spain | Cohort | No | 101 | 2 | 27.9 | 1008.5 | Chorio /HDP/SGA | ANY | 3 | 1 | 3 | 7 |
| Boo 2006 (3) | Malaysia | Cohort | Yes | 60 | 1 | 29.0 | 1115.8 | HDP | INDO | 3 | 2 | 3 | 8 |
| Bravo 2011 (4) | Spain | Cohort | Yes | 90 | 1 | 28.4 | 1066.7 | HDP/SGA | ANY | 4 | 2 | 3 | 9 |
| Brooks 2005 (5) | Australia | Cohort | No | 98 | 1 | 26.0 | 900.5 | SGA | INDO | 3 | 2 | 3 | 8 |
| Dani 2008 (6) | Italy | Cohort | Yes | 26 | 1 | 25.6 | 786.7 | HDP | IBU | 4 | 2 | 3 | 9 |
| Dix 2016 (7) | Netherlands | Cohort | Yes | 76 | 1 | 29.1 | 1312.4 | SGA | INDO | 3 | 1 | 3 | 7 |
| Engeseth 2020 (8) | Germany | Cohort | Yes | 91 | 1 | 26.7 | 865.4 | SGA | INDO | 3 | 1 | 3 | 7 |
| Godambe 2006 (9) | Canada | Cohort | No | 107 | 1 | 26.3 | 887.4 | Chorio/HDP | INDO | 3 | 2 | 3 | 8 |
| Härkin 2018 (10) | Finland | Cohort | No | 1132 | Multi | 29.1 | 1243.0 | SGA | ANY | 4 | 1 | 3 | 8 |
| Hsu 2010 (11) | Taiwan | Cohort | Yes | 31 | 1 | 30.0 | 1396.0 | HDP/SGA | INDO | 2 | 2 | 3 | 7 |
| Hsu 2019 (12) | Taiwan | Cohort | No | 18 | 1 | 27.6 | 1075.0 | SGA | IBU | 3 | 1 | 3 | 7 |
| Itabashi 2003 (13) | Japan | Cohort | No | 2508 | Multi | 28.3 | 1042.0 | HDP | INDO | 4 | 2 | 2 | 8 |
| Kim 2010 (14) | Korea | Cohort | No | 78 | 1 | 26.4 | 742.0 | Chorio/SGA | INDO | 2 | 2 | 3 | 7 |
| Lee 2020 (15) | Korea | Cohort | No | 1063 | Multi | 27.5 | 996.9 | Chorio/HDP/SGA | ANY | 4 | 2 | 3 | 9 |
| Louis 2018 (16) | Canada | Cohort | No | 98 | 1 | 25.4 | 772.0 | SGA | INDO | 3 | 1 | 3 | 7 |
| Madan 2009 (17) | USA | Cohort | Yes | 2435 | Multi | 25.2 | 736.2 | SGA | INDO | 4 | 2 | 2 | 8 |
| Mirea 2012 (18) | Canada | Cohort | No | 2652 | 22 | 26.6 | ND | Chorio/HDP/SGA | INDO | 3 | 2 | 3 | 8 |
| Mitra 2015 (19) | Canada | Cohort | No | 77 | 1 | 25.1 | 840.7 | Chorio | INDO | 3 | 2 | 2 | 7 |
| Mydam 2019 (20) | USA | Cohort | No | 91 | 1 | 25.4 | 827.9 | Chorio | INDO | 3 | 2 | 2 | 7 |
| Oh 2020 (21) | Korea | Cohort | No | 92 | 1 | 27.0 | 939.9 | Chorio/HDP | IBU | 3 | 2 | 3 | 8 |
| Pees 2010 (22) | Germany | Cohort | Yes | 15 | 1 | 26.2 | 853.8 | Chorio | IBU | 2 | 2 | 3 | 7 |
| Rooney 2019 (23) | USA | Cohort | Yes | 133 | 1 | 26.0 | ND | Chorio/HDP/SGA | INDO | 2 | 2 | 3 | 7 |
| Sadeck 2014 (24) | Brazil | Cohort | No | 307 | 16 | 27.3 | 787.1 | SGA | ANY | 3 | 1 | 3 | 7 |
| Sallmon 2018 (25) | Germany | Cohort | No | 471 | 2 | 26.0 | 867.0 | HDP | ANY | 3 | 2 | 3 | 8 |
| Seon 2013 (26) | Korea | Cohort | No | 58 | 1 | 27.1 | 946.0 | Chorio/HDP | IBU | 3 | 1 | 2 | 6 |
| Shah 2011 (27) | USA | Cohort | Yes | 397 | 1 | 25.9 | 828.2 | Chorio /HDP/SGA | INDO | 3 | 2 | 3 | 8 |
| Uchiyama 2011 (28) | Japan | Cohort | No | 57 | 1 | 28.3 | 1040.6 | HDP | INDO | 2 | 2 | 3 | 7 |
| Weiss 1995 (29) | USA | Cohort | Yes | 77 | 1 | 27.8 | 1076.6 | Chorio/HDP | INDO | 3 | 1 | 3 | 7 |
| Yang 2008 (30) | Singapore | Cohort | No | 40 | 1 | 26.3 | 807.3 | SGA | INDO | 3 | 1 | 3 | 7 |

BW: birth weight, Chorio: chorioamnionitis; GA: gestational age, HDP: hypertensive disorders of pregnancy; IBU: ibuprofen, INDO: indomethacin, SGA: small for gestational age.
